# Improving Differentiation of Crohn’s Disease and Ulcerative Colitis Proteomes through Protein-Wide Association Study Feature Selection in Machine Learning

**DOI:** 10.1101/2024.11.13.24316854

**Authors:** Mark G. Gorelik, Aaron J. Gorelik, Skye R.S. Fishbein, Tara Fehlmann, Parakkal Deepak, Ryan Bogdan, Gautam Dantas, Umang Jain, SPARC IBD Investigators

## Abstract

**Background and Aims:** Diagnostic differentiation between Crohn’s disease (CD) and ulcerative colitis (UC) is crucial for timely and suitable therapeutic measures. The current gold standard for differentiating between CD and UC involves endoscopy and histology, which are invasive and costly. We aimed to identify blood plasma proteomic signatures using a Protein-Wide Association Study (PWAS) approach to differentiate CD from UC and evaluate the efficacy of these signatures as features in machine learning (ML) classifiers.

**Methods:** Among participants (n=1,106; n_CD_=636; n_UC_=470) of the Study of a Prospective Adult Research Cohort with IBD (SPARC), plasma protein (n=2,920) levels were estimated using Olink proteomics. A PWAS with Bonferroni correction for multiple testing was used to identify proteins associated with disease states after controlling for age, sex, and disease severity. ML classifiers examined the diagnostic utility of these models. Feature importance was determined via SHapley Additive exPlanations (SHAP) analysis.

**Results:** Thirteen proteins which were significantly differentially abundant in CD vs UC (all |β|s > 0.22, all adjusted p values < 8.42E-06). Random forest models of proteins differentiated between CD and UC with models trained only on PWAS identified proteins (Average ROC-AUC 0.73) outperforming models trained of the full proteome (Average ROC-AUC 0.62). SHAP analysis revealed that Granzyme B, insulin-like peptide 5 (INSL5), and interleukin-12 subunit beta (IL-12B) were the most important features.

**Conclusions:** Our findings demonstrate that PWAS-based feature selection approaches are a powerful method to identify features in complex, noisy datasets. Importantly, we have identified novel peptide based biomarkers such as INSL5, that can be potentially used to complement existing strategies to differentiate between CD and UC.

## INTRODUCTION

Inflammatory bowel diseases (IBD) are chronic relapsing and remitting inflammatory disorders of the gastrointestinal tract. They affect more than 6 million people worldwide, and in the United States alone more than 70,000 cases of IBD are diagnosed each year^2,3^. Patients with IBD experience markedly decreased quality of life, high disease- and treatment-related morbidity, and often endure complications requiring hospitalizations and surgeries^4–8^. IBD is generally subtyped as either Crohn’s disease (CD) or ulcerative colitis (UC), with each differing in the areas of manifestation and the resulting sequela^9–11^. Specifically, CD can affect any region of the gastrointestinal (GI) tract and generally presents with transmural inflammation, while UC is restricted to the colon and is characterized by mucosal ulceration^9–11^. While CD and UC present distinct clinical complications, CD’s ability to affect any region of the GI tract, including regions affected by UC, makes discriminating between them challenging^12–14^.

As each disease requires distinct therapeutic strategies, being able to accurately and efficiently differentiate CD from UC has significant consequences for clinical care. For example, surgery is not a definitive cure for CD and can result in further complications^15–19^. Current practices rely on endoscopy to discriminate CD from UC; however, endoscopy is invasive, expensive, and carries significant risk to the patient^20^. To complement endoscopic procedures, blood and fecal markers are often used; however, none of these tests have proven sufficient to enable the differentiation of CD and UC^21–27^. For instance, serum antibodies against *Saccharomyces cerevisiae* (ASCA) and bacterial antigens have limited accuracy and suffer from low sensitivity, rendering these tests relatively nonspecific to subtype IBD^21–24^. Other markers such as fecal calprotectin and Lipocalin-2 can identify inflammatory status but do not enable differentiation between CD and UC^25–27^. Given the rising prevalence of IBD worldwide, its high morbidity, and its substantial negative impact on quality of life, there is an urgent need for diagnostic tools that enable early differentiation of CD from UC, and are easier to use, non-invasive, and less costly than those currently available^28^.

Advanced proteomics technologies offer novel avenues for comprehending pathophysiological mechanisms and pinpointing potential clinical biomarkers in complex diseases. Recent breakthroughs, exemplified by the Olink platform, have revealed novel protein biomarkers for multiple diseases in blood and plasma ^29,30,31^, ensuring heightened sensitivity, precision, and specificity, while also requiring minimal sample volumes. The output data of the Olink platform can be then applied as features for machine learning (ML)-based classification analysis^32^. Unfortunately, even with these technological advancements, omics data is still often affected by the “curse of dimensionality”^33^, where the number of features captured far exceeds the number of samples which can result in models fitting to spurious patterns^33^. ML models trained on high dimensionality data may fail to generalize to real world data unless the sample size is sufficiently large enough (normally at least 5 samples per feature^34^) to separate signal versus noise^33^. However, generating such large omics datasets can be both costly and time consuming. To mitigate the “curse of dimensionality” without the costly and time-consuming process of generating large omics datasets, feature selection methods are often used to identify informative features in high dimensionality datasets before model training^33,35^. In particular, GWAS (Genome-Wide Association Study) and PheWAS (Phenome-Wide Association Study) approaches have proven to be extremely effective for feature selection^35^. Here we leverage a PWAS (Protein-Wide Association Study)-based approach to identify informative features in a high dimensionality Olink proteomics dataset from IBD patients. This approach identified 13 proteins which distinguish CD from UC using plasma samples.

## MATERIALS & METHODS

### Participants and Sample Collection

The Study of Prospective Adult Research Cohort of IBD (SPARC IBD) is an ongoing longitudinal cohort study of patients with IBD recruited from 17 academic medical centers across the United States^1^. Plasma samples used in this study were obtained from N = 1106 individuals (n_CD_ = 636; n_UC_ =470).

Demographic, disease-related, and patient-reported data were collected during the following visits: 1) during routine GI office visits (2016-2021), 2) quarterly by sending surveys to patients, and 3) before a scheduled colonoscopy. All collections generated highly structured electronic case report forms (eCRF). Bio-samples of each respective patient’s blood and stool were collected at enrolment and at the time of each patient’s colonoscopy. Further, blood samples were collected if a patient or provider reported key medication changes. Initially collected samples were used in this study. Clinical data is transferred from sites on a periodic basis and stored in IBD Plexus, Crohn’s & Colitis Foundation’s exchange platform (see^1^ for details).

### Olink Proteomics, normalization, and filtering

Plasma was purified from blood and stored in EDTA. Proteins within plasma were estimated using Olink Explore 384 panels (i.e., Cardiometabolic, Cardiometabolic II, Inflammation, Inflammation II, Neurology, Neurology II, Oncology, Oncology II panels; Olink Proteomics) Protein levels were estimated as Olink’s arbitrary units, Normalized Protein eXpression (NPX) values on a log_2_ scale. NPX values which did not pass the following quality control metrics were filtered out: 1) at least 500 counts per specific combination of sample and assay, 2) the deviation from the median value of the incubation- and amplification controls for each individual sample did not exceed +/-0.3 NPX for either of the internal controls, and 3) the deviation of the median of the negative controls must be ≤5 standard deviations from the set predefined manufacturer value. Samples across plates were normalized via the intensity normalization method. The following Explore 384 assays did not meet Olink’s batch release quality control criteria and are therefore not included in this study: KNG1 (Inflammation II), TNFSF9 (Inflammation II), TOM1L2 (Neurology II), SMAD1 (Oncology), and ARHGAP25 (Oncology).

### Statistical Analyses

#### Protein Wide Association Study (PWAS)

A Protein Wide Association Study (PWAS) was performed on all proteins passing quality control described above (n=2,920) using the glmer function in the lme4 package^36^ as previously described in phenome-wide association studies^37,38^. CD/UC disease status was regressed on each individual protein in a mixed effects logistic regression with age and sex as fixed effects covariates and disease activity (Simple Crohn’s Disease Activity Index for CD and 6-point Mayo Score for UC^1^) was treated as a random effect. To adjust for multiple testing, a Bonferroni-corrected proteome wide significance threshold was used (0.05/2, 920 = 0.0000171 alpha level).

#### Principle Component Analysis (PCA)

Principal component analysis (PCA) was initially performed incorporating the measured values of all proteins and just proteins identified via the PWAS analysis. Analysis of Similarities (ANOSIM) was performed using the ANOSIM function in the vegan package^39^.

#### Machine Learning Methods

Using Scikit learn based implementations of random forests we tested the following feature sets: All proteomics features and patient features (Age, Sex, Disease Severity), proteomics features which passed the Bonferroni cutoff and patient features, and just proteomics features. Of the samples, 20% were reserved for a holdout validation dataset which was also used for SHAP value analysis^40^. The remaining 80% of the data was split into a train/test split (70/30) and cross validated 30 times.

The following packages and versions were used for analysis in R v4.3.2: ibdplexus (0.1.0), tidyverse (1.3.1), stringr (1.5.1), readxl (1.4.3), OlinkAnalyze (3.7.0), data.table (1.15.0), lmerTest (3.1), lme4 (1.1)^36^, readxl (1.4.3), dplyr (1.1.4), ggplot2 (3.4.3), reshape2 (1.4.4), ggrepel (0.9.5), forcats (1.0.0), ggsci (3.0.0), RColorBrewer (1.1.3), optimx (2023-10.21), minqa (1.2.6), dfoptim (2023.1.0), survey (4.2.1), scales (1.3.0), ggnewscale (0.4.10), ggpubr (0.6.0), gplots (3.1.31), psych (2.4.1), MuMIn (1.47.5), vegan (2-6-6.1), NatParksPalettes (0.2.0), and ggfortify (0.4.16).

The following packages and versions were used for analysis in Python v3.10.9: pandas^41^(1.5.3), sklearn^42,43^ (1.3.2), numpy^44^ (1.23.5), and shap^40^(0.43.0).

#### Formulas

TP: True Positive

TN: False Negative

FP: False Positive

FN: False Negative

Accuracy = (TP+FN)/(TP+FP+TN+FN)

Sensitivity=TP/(TP+FN) Specificity=TN/(TN+FP)

## RESULTS

### Subject Characteristics

Details of the subjects’ characteristics are shown in Table 1. A total of 1,106 individuals (CD n = 636; UC n=470) from 17 medical centers were included. CD patients were on average, 42.26 years old and 62% of the CD patients were female. UC patients were on average 43.98 years old and 50.95% of the total UC patients were female.

**Table 1.**
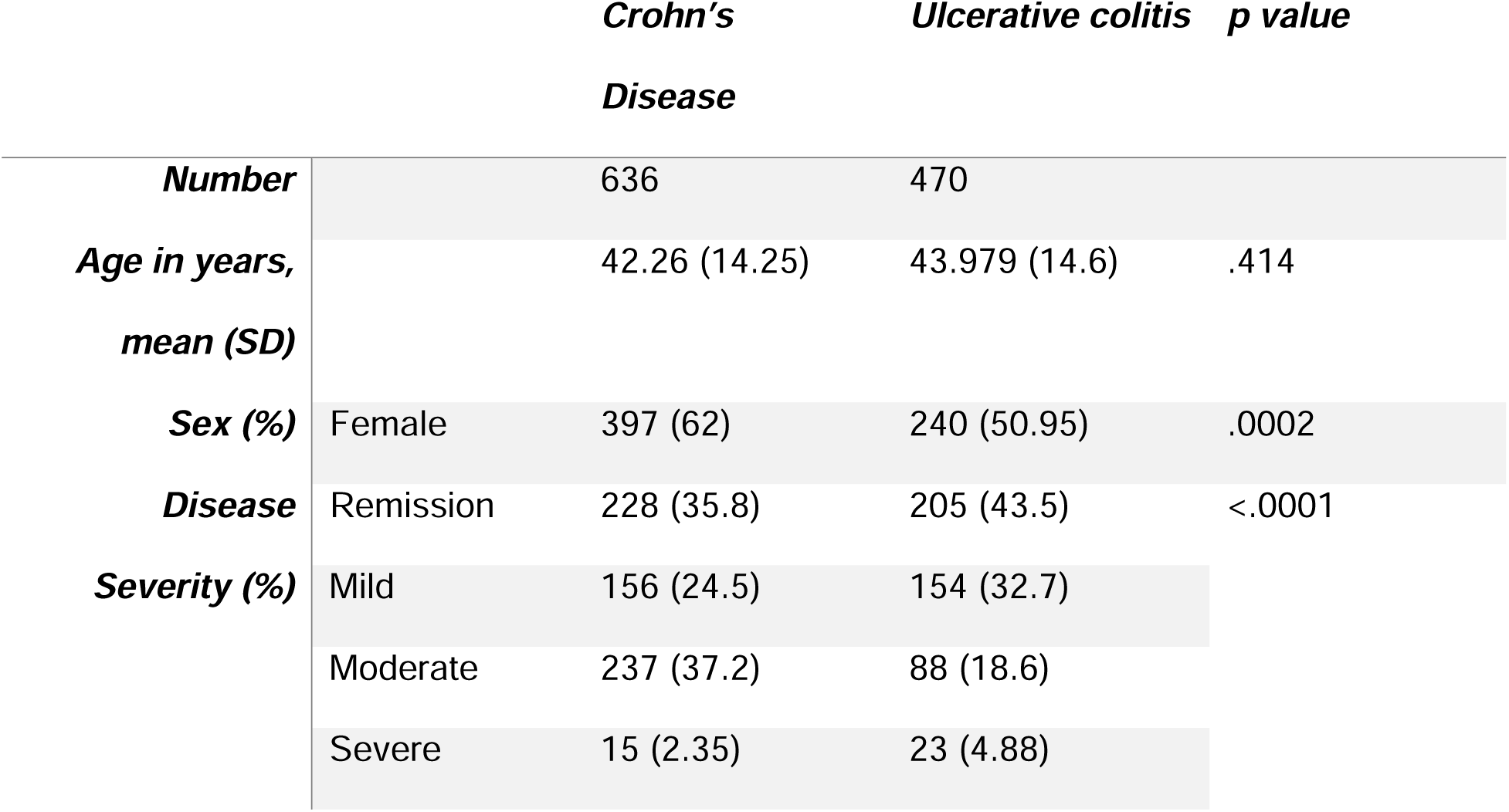
Cohort Breakdown.

### Specific proteins differentiate the proteomic profiles of Crohn’s disease and Ulcerative colitis

The studied plasma proteome dataset includes measures of 2,920 protein levels across 1,106 patients from the Crohn’s and Colitis Foundation dataset^1^ **(Table 1**, **Fig 1)**. We initially ran a principal component analysis (PCA) (**Fig 2A**) and an analysis of similarities (ANOSIM) which revealed that the global proteomes of CD and UC do not differ (p=0.21, R=0.004). We then conducted a Protein Wide Association Study (PWAS) analysis adapted from previous study ^38,45^ to filter for proteins measured at significantly different levels between CD and UC. Age and sex were included as fixed effects with disease severity as random effect. The PWAS-based approach identified thirteen proteins that were significant after Bonferroni correction **(Table 2**, **Fig 2B, Table S1)**. Five of these proteins, INSL5, IL12B, IL12AB, HRG, and LY96 were more abundant in CD relative to UC; in contrast, eight proteins, FGF19, EPCAM, NOS2, GPA33, GUC2A, GRAB, FGFR4, MMP10 were more abundant in UC relative to CD (**Fig S1**). Performing an ANOSIM test and PCA analysis (**Fig 2C**) on the proteins identified via PWAS after multiple test correction revealed that there was significant a difference between the CD and UC cohorts (p=0.001, R=0.1247).

**Figure 1.**
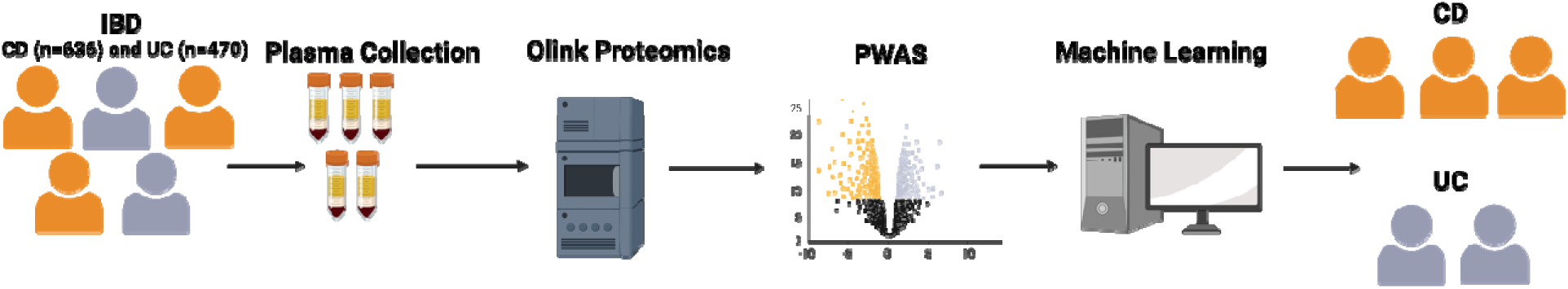
Sample processing and analysis pipeline. Blood plasma samples were collected and processed as described in the methods and materials. Differentially abundant proteins were identified in the PWAS analysis. Protein abundance was used as features for the machine learning models to classify CD from UC.

**Figure 2.**
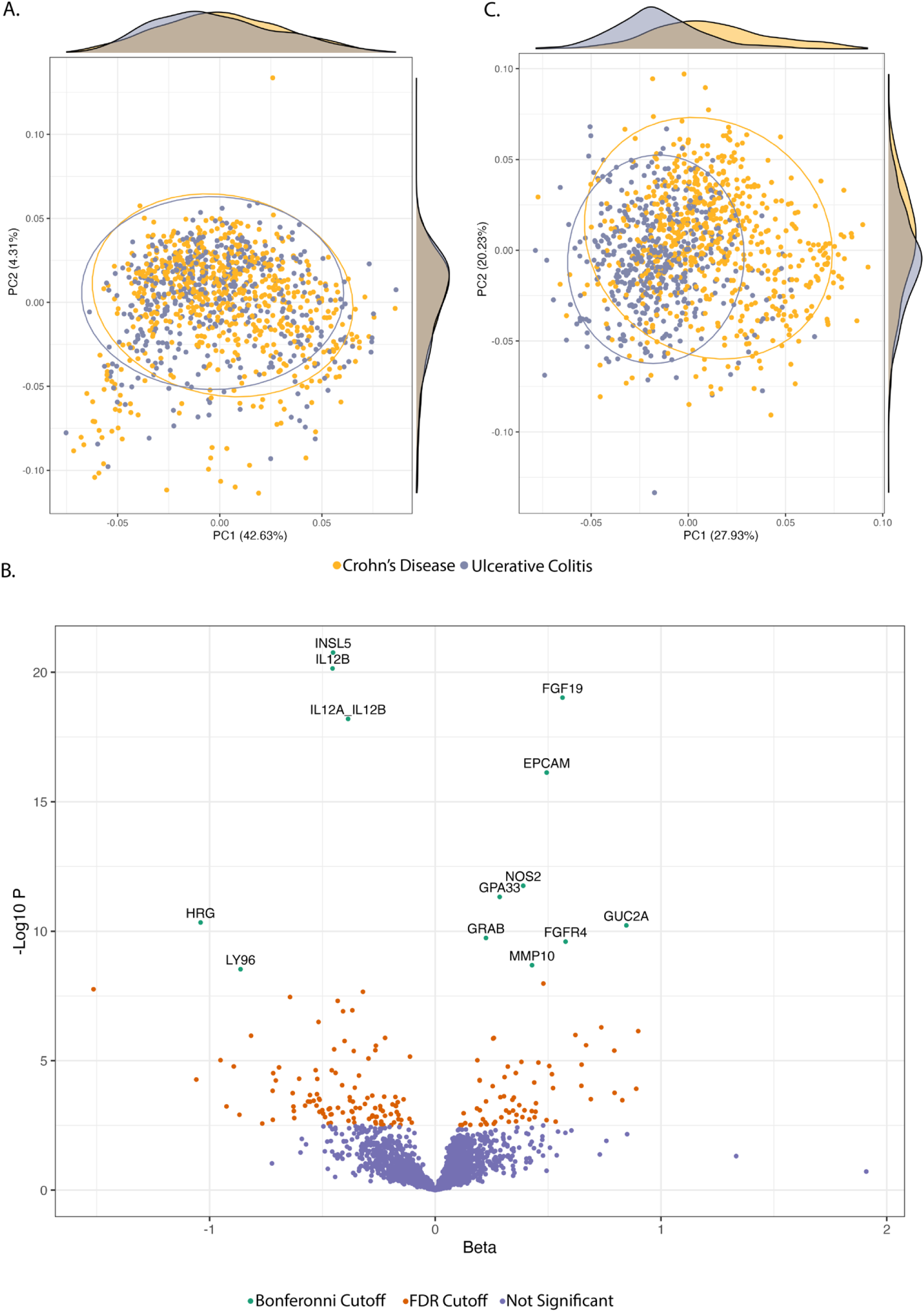
PWAS analysis enables separation of the proteomic profiles of Ulcerative colitis and Crohn’s disease. A) Principal Component Analysis (PCA) of the global proteomics profiles of Crohn’s disease and Ulcerative colitis. B) Volcano plot where the x axis is the calculated beta and the y axis is the negative log_10_ of the unadjusted p-value; green and labeled points had a Bonferroni adjusted p-value of less than 0.0000171 (used in Fig 1B), orange points had an FDR adjusted p value of less than .05, and purple points represent proteins with a p-value > .05. Negative beta values are associated with Crohn’s disease and positive beta values are associated with Ulcerative colitis. C) PCA of the proteomics profile identified by the PWAS analysis. Ellipses represent 95% confidence bounds around group centroids.

**Table 2.**
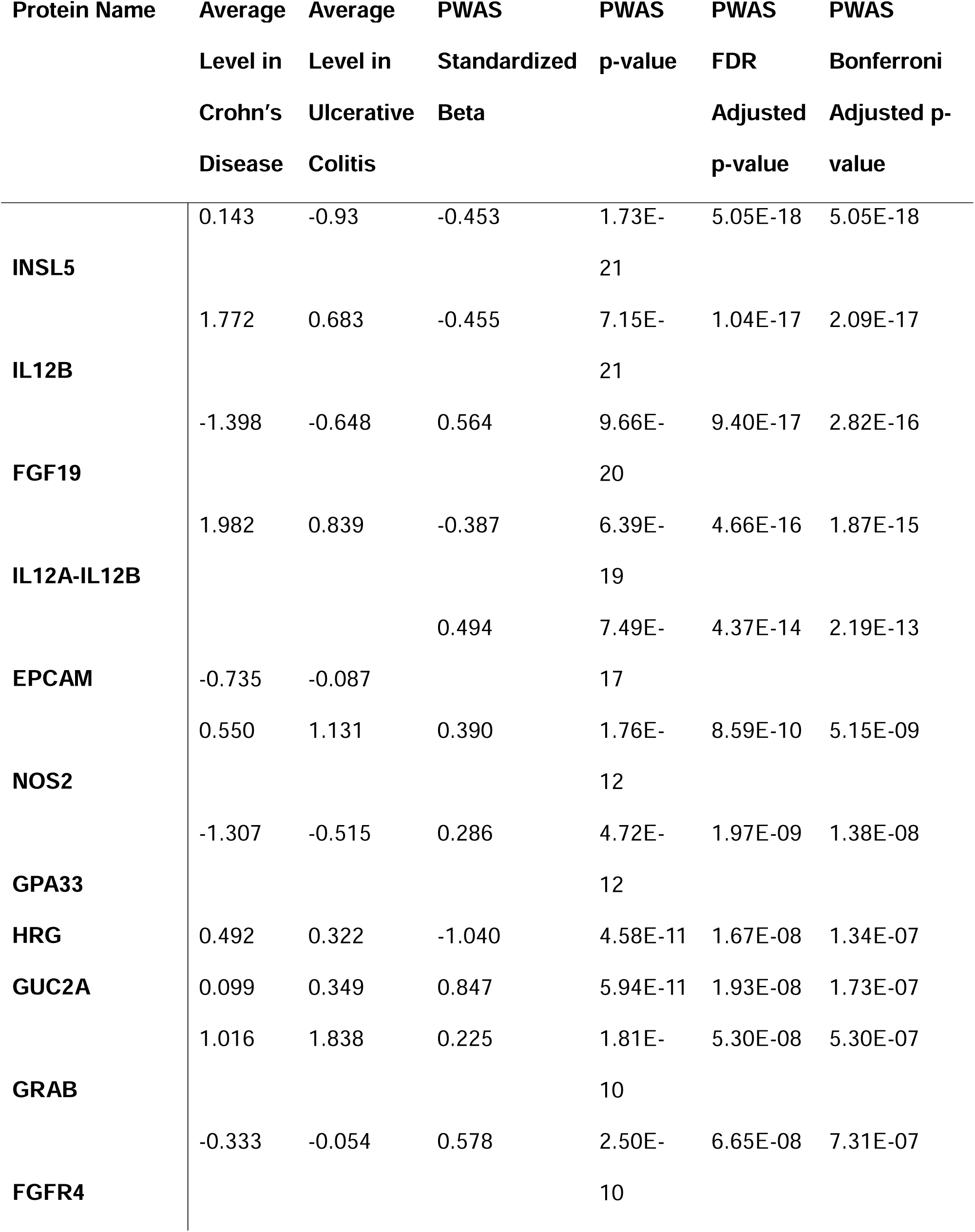
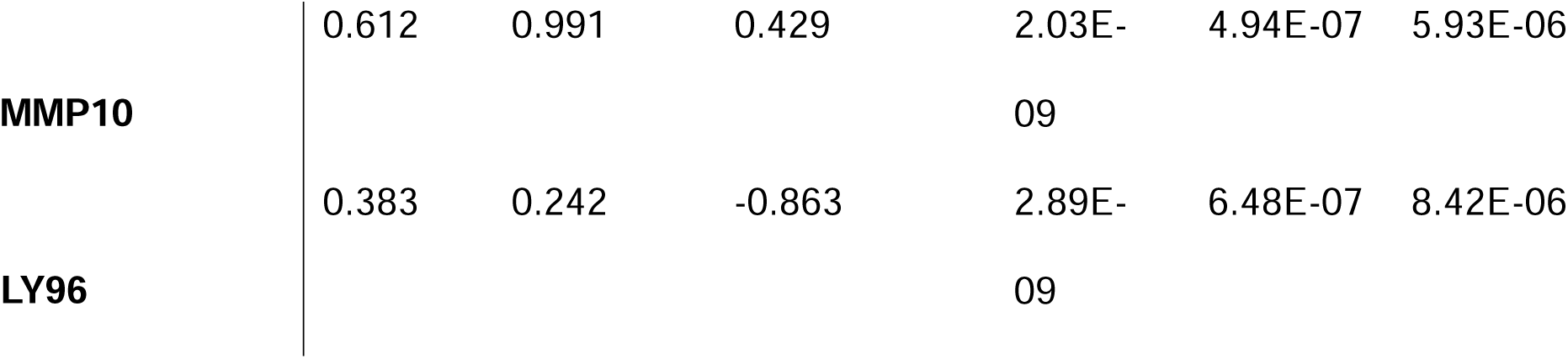
PWAS Results.

### Grouping Specific proteins improve prediction of Crohn’s disease and Ulcerative Colitis

We aimed to determine whether the PWAS identified proteomic features could lead to improved differentiation of CD and UC via ML classification. To do this we trained random forest models on feature sets composed of different combinations of proteomic features and patient features (age, sex, disease severity). We generated three features sets which contained the following: 1) the entire proteome and patient features (“Full Feature Set”), 2) subset of proteomic features which only included the thirteen significant proteins (e.g., INSL5) identified by the PWAS and patient features (referred to as “PWAS and Patient Features”), and 3) just the thirteen significant proteins identified by the PWAS without patient features (“PWAS Features”). Models trained on the “Full Feature Set” had significantly higher specificity, but significantly lower accuracy, sensitivity, and ROC-AUC scores compared to the other two feature sets **(Fig 3A-D**, **Table 3**). Models trained on “PWAS and Patient Features” and just “PWAS Features” were significantly more accurate and sensitive than models trained on the “Full Feature Set,” they did not differ significantly in performance from each other (**Fig 3A-D**). Next, we interrogated the ML models using SHapley Additive exPlanations (SHAP) analysis, which can infer feature importance, to determine if patient features were important for model performance. Interestingly, SHAP feature importance analysis suggested patient features were not informative, where Age, Disease Severity, and Sex were the ranked as the three least important features in the “PWAS and Patient Features” model suggesting that the patient features contributed the least to model performance **(Fig 2B, 4AB, S1)**. Notably, interrogating the ML models revealed that the three most important features including granzyme B, Insulin-like peptide 5 (INSL5), and Interleukin 12B (IL12B) were conserved between models. **(Fig 4AB)**. This demonstrates that PWAS-based approaches act as a filter for identifying more informative features which in turn improve prediction performance.

**Figure 3.**
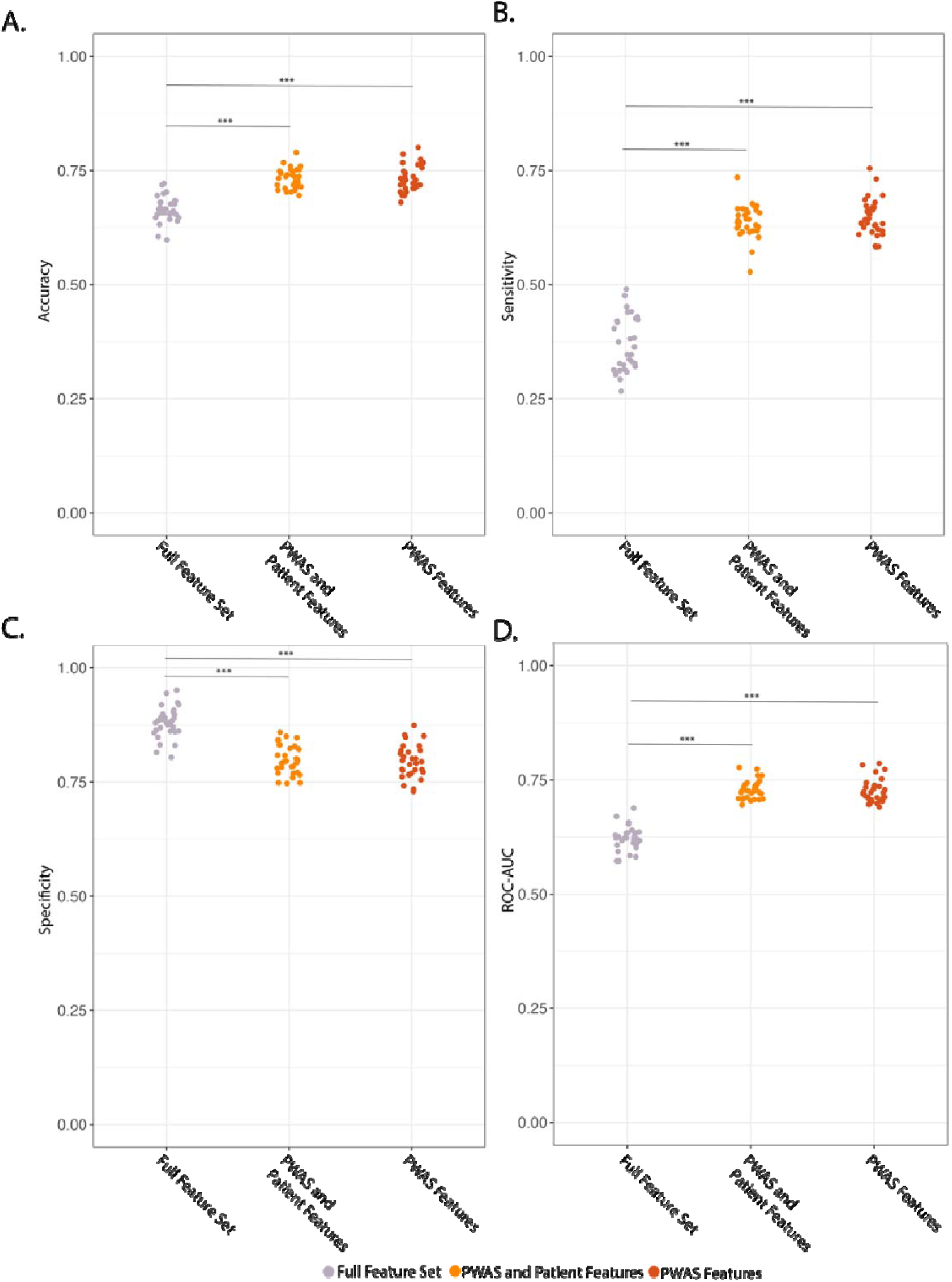
Specific proteins improve machine learning based differentiation of CD and UC. A) Effect of feature set on model accuracy. B) Effect of feature set on model sensitivity. C) Effect of feature set on model specificity. D) Effect of feature set on model ROC-AUC. ***P<.001, ANOVA with Tukey’s post hoc test.

**Figure 4.**
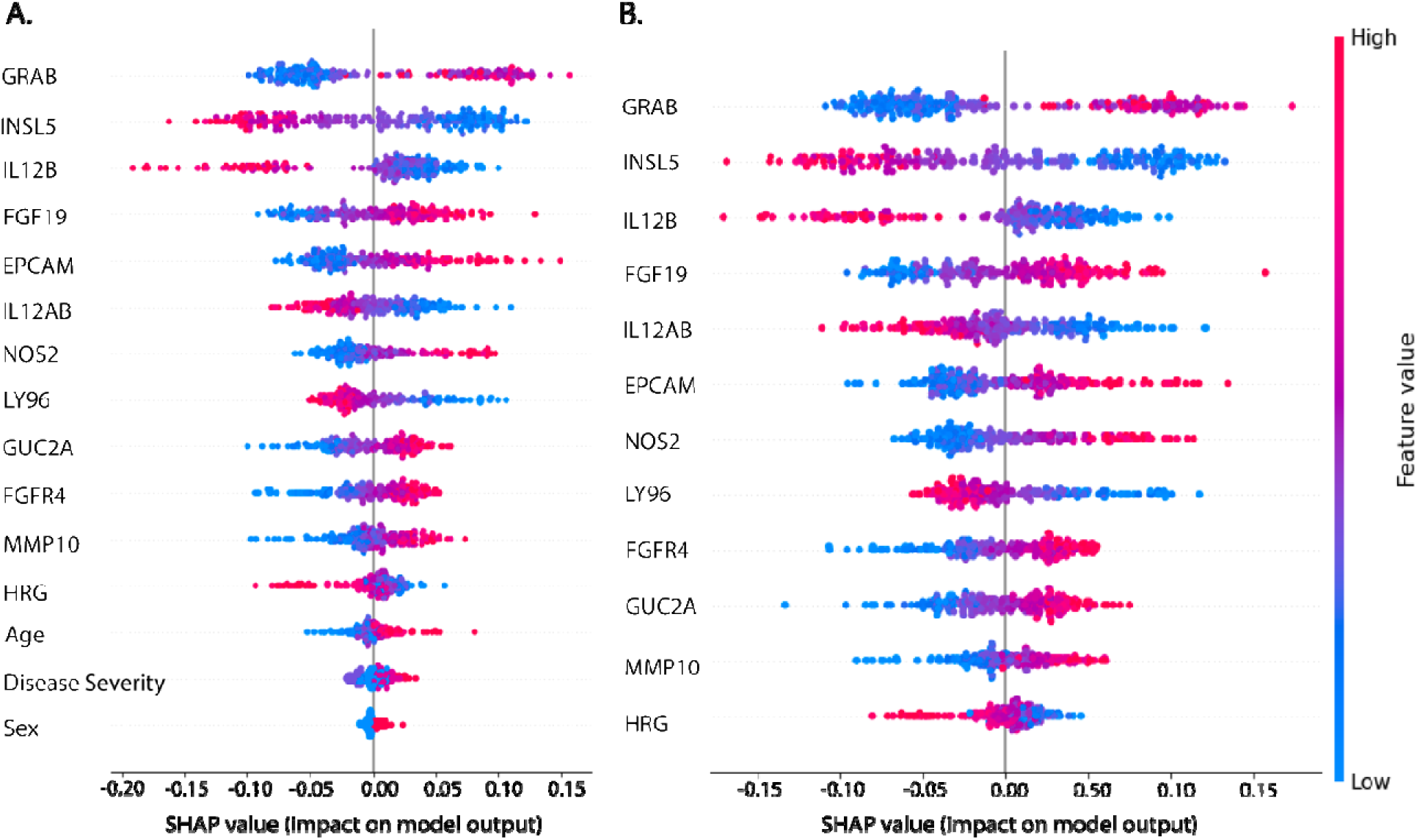
Clinical Features do not improve model performance. A) SHAP beeswarm plot of the validation dataset indicating feature importance in random forest models trained on patient associated features (Age, Sex, Disease Severity) and the thirteen proteins which are significantly associated with Crohn’s disease and Ulcerative colitis. B) SHAP beeswarm plot of the validation dataset indicating feature importance in random forest models trained on just the thirteen proteins which are significantly associated with Crohn’s disease and ulcerative colitis. Features are sorted in order of predicted importance in a descending manner.

**Table 3.**
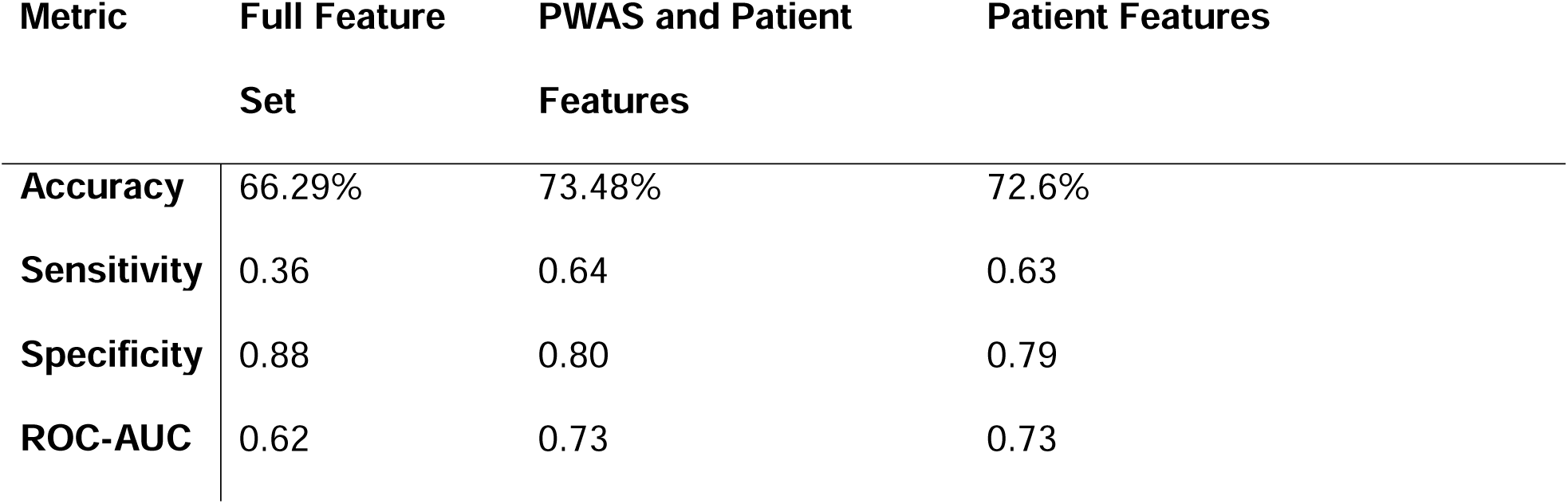
Average Machine Learning Model Results.

| **Metric** | **Full Feature Set** | **PWAS and Patient Features** | **Patient Features** |
| --- | --- | --- | --- |
| **Accuracy** | 66.29% | 73.48% | 72.6% |
| **Sensitivity** | 0.36 | 0.64 | 0.63 |
| **Specificity** | 0.88 | 0.80 | 0.79 |
| **ROC-AUC** | 0.62 | 0.73 | 0.73 |

## DISCUSSION

Accurately classifying subtypes of IBD poses a significant clinical challenge. Identification of noninvasive biomarkers that can increase the accuracy of diagnosing and subtyping IBD is a major unmet need. There has been a growing interest in using proteomics to identify new biomarkers for the differentiation of IBD; however, these studies have been limited by the number of proteins measured^46,47^. Here, we used a highly sensitive proximity extension assay and measured 2920 proteins in the plasma of IBD patients. Protein wide association analysis with age and sex as fixed effects identified 13 proteins that are significantly different between CD and UC. Further, using used multiple feature sets in random forest models, we discovered that PWAS identified proteins could distinguish between CD and UC with high accuracy and sensitivity. Taken together, we have identified a novel set of proteins in blood that can potentially complement other existing biomarkers to accurately subtype IBD.

Machine learning algorithms are increasingly being utilized to analyze medical data to diagnose diseases, predict their severity, and monitor their progression. Recent work on diagnosing IBD using ML approaches has also been successful, achieving high levels of performance^12,48–50^. For instance, supervised learning models on RNA sequencing data enabled CD and UC differentiation^12^. Similarly, deep learning networks have been used on endoscopic images to accurately predict the severity of the disease in IBD^48,49^. Although proteomic datasets have been generated in IBD, the application of ML techniques to analyze such datasets has been limited^47^. Furthermore, previous IBD-focused proteomics datasets have measured smaller panels of proteins^47,51–53^. We used a combined PWAS-based feature selection and ML models on a large dataset of proteins to identify novel signatures that could accurately subtype IBD. Importantly, in contrast to previous studies that have primarily focused on inflammatory markers, we analyzed proteins that are involved in a diverse array of processes including, hormonal regulation, inflammation, cancer, and brain gut axis. Our findings suggest that PWAS based ML approaches could improve subtyping of IBD patients.

Several proteins in our cohort have been validated by other IBD studies focused on differentiating CD from UC. A study by Bourgonje et al. also employed a proximity extension assay (Olink) and measured 92 proteins identifying FGF19, IL12B, and MMP10 to be differentially abundant between CD and UC^53^. In another study by Di Narzo et al., the authors used a SOMAmer-based capture array to measure protein levels in plasma (n=244) and discovered that Granzyme B, FGF19, and MMP10 were downregulated in CD relative to UC, mirroring our results^52^. Importantly, our findings combined with others have identified FGF19 and MMP10 as consistent plasma-based biomarkers which can be used differentiate CD from UC^52,53^.

Among the 13 differentially abundant proteins significant in the PWAS after multiple correction, Granzyme B, IL12B, and INSL5 were the most informative for model prediction (**Fig 4A-B**). INSL5, to date, has not been measured in similar proteomics studies focused on IBD^52,53^. Notably, depletion of *INSL5* transcripts in mucosal tissue has been associated with IBD^54^, and our study further implicates the INSL5 peptide as differentially abundant between CD and UC. INSL5 is a peptide hormone that is expressed in the colonic epithelium ^54–56^. Because INSL5 is a microbially regulated molecule, it is possible that UC, but not CD, specific microbes alter its production^57^. Indeed, both bacterial and fungal microbiota are known to be different between UC and CD^58–60^. Another possible reason for the decreased abundance of INSL5 in UC relative to CD is the loss of colonic epithelial cells due to ulceration, a prominent feature of UC^61^. Future studies are needed to elucidate how INSL5 is regulated and the mechanisms by which INSL5 modulates the severity of the disease^54,62^. In addition to INSL5, Granzyme B was also a powerful predictive feature and was elevated in UC. Granzyme B is a serine protease released by lymphocytes which can trigger apoptosis^63,64^. Similar to our findings, Di Narzo et al. identified elevated Granzyme B protein levels in CD compared to UC^52^. Further, levels of Granzyme B have been reported to predict treatment responses in IBD as its levels are significantly lower in responders compared to non-responder populations^65^. While these initial findings are promising, it is unclear how the levels of these targets fluctuate throughout disease specific treatments and subtype. Future studies utilizing longitudinal samples are needed to ascertain its association with IBD subtypes.

Our study has several strengths: (1) We utilized a relatively large sample size with patients from 17 different medical centers; (2) Because the SPARC cohort follows standard guidelines, it allows investigators to maintain consistency in both data and bio-sample collection; (3) Our data analysis controlled for multiple parameters including age and sex; (4) We assessed over 2,900 proteins using the Olink platform, enabling us to capture differences across a wide range. Limitations include: (1) absence of a healthy cohort, (2) a single time point of blood collection, (3) and need for validation in non-North American cohorts. Future studies including a healthy control group and longitudinal data would enable exploration of the complex nature of IBD, focusing on the complex spatial-temporal dynamics of IBD location and flare up. This would add important context for leveraging proteins such as INSL5 whether alone or in combination with other markers to differentiate between CD and UC.

Overall, the results of this study provide evidence that applying a PWAS-based approach to filter for potentially relevant proteins improves ML model predication for differentiation between CD and UC. Importantly, the informative biomarkers identified in our study have not been previously examined in the context of differentiating CD from UC. We speculate that this approach may identify new targets for biomarker research and improve mechanistic understanding of disease states.

## Data Availability

The SPARC IBD data are available upon approved application to Crohns & Colitis Foundation IBD Plexus

## SPARC-IBD Investigators

|  |  |
| --- | --- |
| Baylor College of Medicine | Richa Shukla, MD |
| Baylor University Medical Center | Themistocles Dassopoulos, MD |
| Brigham & Women's Hospital | Scott B. Snapper, MD, PhD Joshua R. Korzenik, MD |
| Indiana University | Matthew Bohm, MD |
| Mayo Clinic | Laura Raffals, MD |
| Medical College of Wisconsin | Poonam Beniwal-Patel, MD |
| NYU Langone Medical Center | David Hudesman, MD |
| Scripps Healthcare | Mazer Ally, MD Gauree Konijeti, MD Rebecca Matro, MD |
| University Gastroenterology | Sheldon Lidofsky, MD |
| University of Alabama | Kirk Russ, MD |
| University of Cincinnati Medical Center | Loren Brook, MD |
| University of Chicago | Joel Pekow, MD |
| University of Maryland | Raymond Cross, MD |
| University of Michigan | Shrinivas Bishu, MD |
| University of Pennsylvania | Meenakshi Bewtra, MD, MPH, PhD James D Lewis, MD, MSCE |
| University of Pittsburgh | Richard Duerr, MD |
| University of Wisconsin | Sumona Saha MD, MS Freddy Caldera DO, MS |
| Vanderbilt University Medical Center | Elizabeth Scoville, MD, MSCI |
| Washington University School of Medicine | Parakkal Deepak, MBBS, MS |

## Funding information

This work is supported by SPARC IBD PLEXUS Grant from Crohn’s & Colitis Foundation and NIH Washington University-DDRCC Grant Number P30 DK052574 to U.J. M.G.G and S.R.F are supported by awards from the Pediatric Gastroenterology Research Training Program (T32 DK077653). A.J.G is supported by NSF (DGE-213989). PD is supported by a Junior Faculty Development Award from the American College of Gastroenterology and IBD Plexus of the Crohn’s & Colitis Foundation.

## Presentation at a meeting

A portion of this study was presented at the 2024 Digestive and Diseases Week (May 18-21) in Washington D.C.

## Acknowledgements

The authors would like to thank Sarah E. Paul for her initial analytical support and Kevin S. Blake of the LGM Scientific Editing Service at the Department of Pathology and Immunology, Washington University School of Medicine for scientific editing support. The authors also thank the staff at The Edison Family Center for Genome Sciences & Systems Biology at the Washington University School of Medicine in St Louis, including E. Martin and B. Koebbe for computational support, and B. Dee, K. Matheny, J. Theodore and K. Page for administrative support. Finally, we would like to thank the members of the Dantas, Bogdan, and Jain labs for helpful general discussions and comments on the manuscript. The results published here are in whole based on data from the Study of a Prospective Adult Research Cohort with IBD (SPARC IBD). SPARC IBD is a component of the Crohn’s & Colitis Foundation’s IBD Plexus data exchange platform. SPARC IBD enrolls patients with an established or new diagnosis of IBD from sites throughout the United States and links data collected from the electronic health record and study specific case report forms. Patients also provide blood, stool and biopsy samples at selected times during follow-up. The design and implementation of the SPARC IBD cohort has been previously described^1^. The SPARC IBD data are available upon approved application to Crohn’s & Colitis Foundation IBD Plexus (https://www.crohnscolitisfoundation.org/ibd-plexus).

## Conflict of Interest

PD: has received research support under a sponsored research agreement unrelated to the data in the paper and/or consulting from AbbVie, Arena Pharmaceuticals, Boehringer Ingelheim, Bristol Myers Squibb, Janssen, Pfizer, Prometheus Biosciences, Takeda Pharmaceuticals, Roche Genentech, Scipher Medicine, Fresenius Kabi, Teva Pharmaceuticals, Landos Pharmaceuticals, Iterative scopes and CorEvitas, LLC. U.J. has received research support from Boehringer Ingelheim.

## Ethics Approval

The study protocol was approved by the Institutional Review Board (IRB) at the University of Pennsylvania which is the single IRB for the SPARC IBD study.

## Author contributions

M.G.G., P.D., R.B., G.D. and U.J designed the study, T.F., P.D., and U.J. collected and acquired data, M.G.G., A.J.G., S. R.S.F., and T.F.analysed data, M.G.G., and U.J wrote the manuscript. All authors approved the manuscript.

## Abbreviations

IBD: Inflammatory bowel diseases
PWAS: Protein-Wide Association Study
CD: Crohn’s disease
UC: Ulcerative colitis

---

**Supplemental figure 1.**
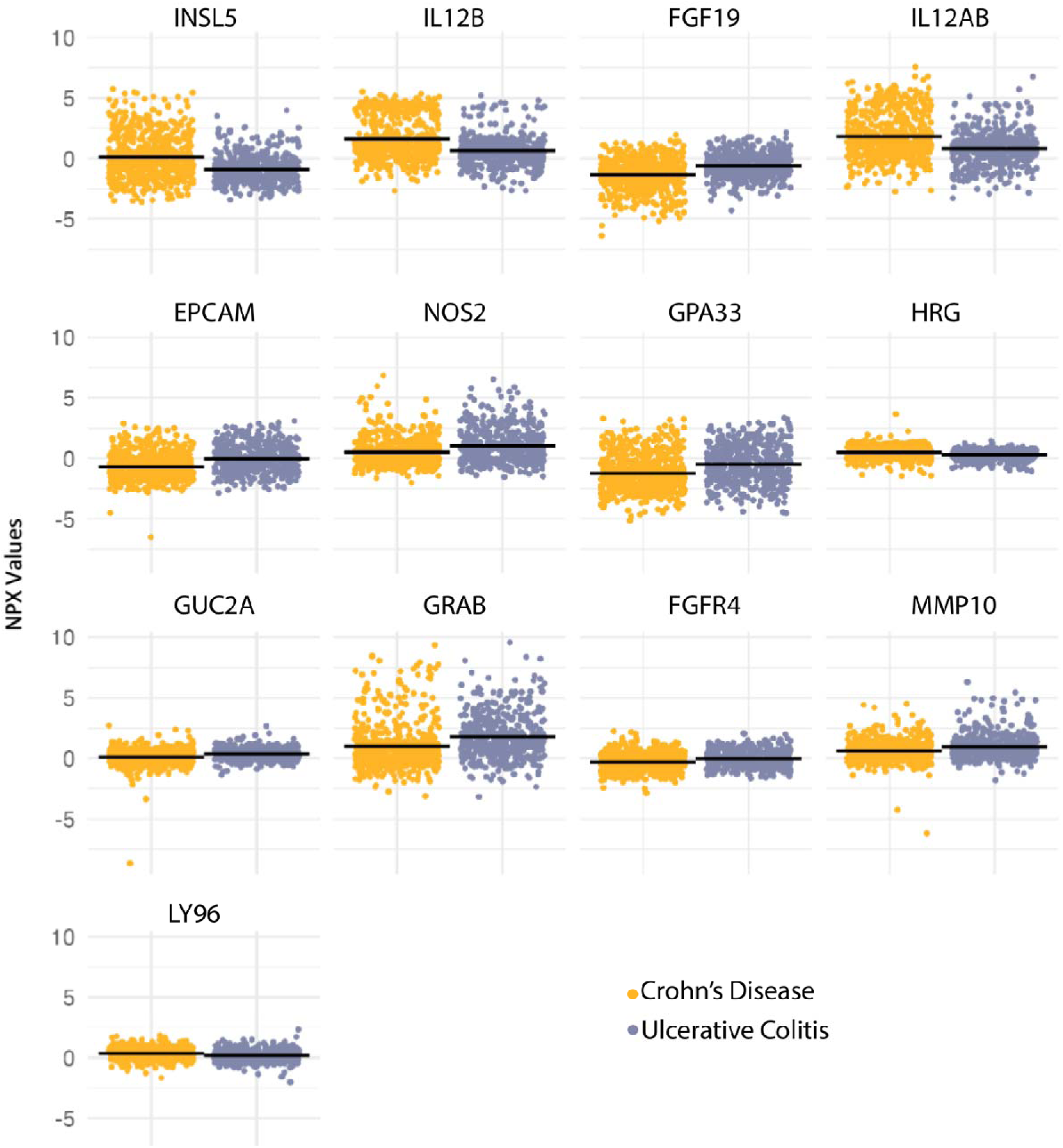
The average NPX values for proteins which were significant after Bonferroni correction (Fig 1B).

## Notes

### Funding Statement

This work is supported by SPARC IBD PLEXUS Grant from CCFA and NIH Washington University-DDRCC Grant Number P30 DK052574 to U.J. M.G.G and S.R.F are supported by awards from the Pediatric Gastroenterology Research Training Program (T32 DK077653). A.J.G is supported by NSF (DGE-213989). PD is supported by a Junior Faculty Development Award from the American College of Gastroenterology and IBD Plexus of the Crohn's & Colitis Foundation

